# Women’s Health Research Funding in Canada across 15 years suggests low funding levels with a narrow focus

**DOI:** 10.1101/2025.04.14.25325826

**Authors:** Laura L Gravelsins, Tallinn FL Splinter, Ahmad Mohammad, Samantha A Blankers, Gabrielle L Desilets, Liisa AM Galea

## Abstract

**Background:** Females have been underrepresented in preclinical and clinical research. Research on females is important for conditions that directly affect women, disproportionately impact women, and manifest differently in women. Sex and gender mandates were introduced, in part, to increase women’s health research. This study aimed to understand how much of women’s health research is being funded in open grant competitions in Canada that fall under the top burden and/or death of disease for women globally.

**Methods:** Publicly available funded Canadian Institute of Health Research (CIHR) project grant abstracts from 2009-2023 were coded for the mention of female-specific research to assess what percentage of grant abstracts focused on the top 11 areas of global disease burden and/or death that disproportionately affect females. We also examined changes from 2020 to 2023 in the representation of grant abstracts that mentioned sex, gender, or two-spirit, lesbian, gay, bisexual, trans, queer, intersex (2S/LGBTQI).

**Results:** The percentage of abstracts mentioning sex or gender doubled whereas the percentage of abstracts mentioning 2S/LGBTQI quadrupled from 2020 to 2023, but remained at under 10% of overall funded abstracts. In contrast, female-specific research representation remained at ~7% of all research. Under 5% of the total funded grant abstracts mentioned studying one of the top 11 global burdens of disease and/or death for women over 15 years. Of the 681 female-specific grants, cancer research accounted for 32% of funding (or 2.09% of overall grants), whereas the other top 10 collectively accounted for 38% of female-specific funding (or 2.45% overall) across 15 years. The percentage of overall funding towards understanding female-specific contributions to cardiovascular disease was 0.83% followed by diabetes (0.41%), HIV/AIDS (0.4%), depression (0.35%), anxiety (0.16%), musculoskeletal (0.12%), dementia (0.08%), respiratory (0.08%), headache (0.01%) and low back pain (0.01%).

**Conclusions:** Research acknowledging the sex and gender population in CIHR abstracts is increasing but remains at under 10% while the percentage of funding for women’s health remains unchanged at 7% of funded grants across 15 years.

**Highlights:**

- From 2020 to 2023, funded grant abstracts that mentioned sex, gender or 2S/LGBTQI populations doubled or quadrupled. Across the same time period, funded grant abstracts that mentioned female-specific research increased by less than a percentage.
- Women’s health research accounts for 7% of all funded grant abstracts and increases were not observed over 15 years.
- Over 15 years, only 4.5% of funded grants examined the 11 causes of global disease burden and/or death that disproportionately affect females using female-specific populations.
- Of these female-specific grants, cancer (breast and gynecological) accounted for over 32% of all the female-specific funded grants whereas headache disorders and depression/anxiety accounted for 0.15% and 6.9% respectively across 15 years. Given the significant burden of these diseases, greater efforts are needed to expand the breadth of women’s health research.

**Plain English Summary:** Women’s health research has historically been underrepresented and underfunded. Here, we examined 15 years of funding data from the Canadian Institute of Health Research (CIHR), Canada’s major funder of medical research, to understand what type of women’s health research is being funded using the top 11 burdens of disease or death globally for women. From 2020 to 2023, the percentage of abstracts mentioning sex or gender doubled, whereas those mentioning 2S/LGBTQI populations quadrupled, yet still remained under 10% of all funded research. In contrast, across the same time period, women’s health research representation increased by less than a percentage and remained at ~7% of all funded research. We also examined the amount of funded research dedicated to the top 11 global burdens of disease and/or death that affect women across 15 years (from 2009-2023). Across these 15 years, we observed that cancer accounted for 2.09% of all funded research and received approximately the same amount of research representation as the 10 other global burdens of disease and death combined (2.45% of all funded research). Despite CIHR’s mandates to encourage the integration of sex and gender into research, sex and gender representation is low and mandates did not increase women’s health research. More efforts and support beyond sex and gender mandates are needed to increase and diversify women’s health research to achieve personalized medicine and close the women’s health gap.

## Background

Women experience delays in diagnoses, and a greater percentage of adverse effects towards new therapeutics than men (1,2). Across nearly 1300 of the same diseases and health conditions, females are diagnosed on average 2 years later than males (1), and women spend 25% more time in poor health and with disability compared to men (3). These disparities stem, in part, from a lack of research in women’s health (4,5). The paucity of women’s health research (i.e. research that focuses on individuals with female sex or who identify as women) may be due to misconceptions that include the belief that human males and females are similar in their physiological responses to treatments and disease endophenotypes. However, dramatic sex differences are seen in gene expression and physiological responses to drugs and disease - often in opposing directions - indicating that basic physiological mechanisms differ between the sexes including disorders that represent the top global burdens of disease and death (6–10). Indeed, the top global burdens of death in women have been identified as cardiovascular diseases (CVDs), diabetes, cancers, dementias, and respiratory diseases (8). In addition to these disorders, the top global burdens of disease in women have identified depressive disorders, headache disorders, anxiety, musculoskeletal disorders, HIV/AIDS, and low back pain (7). Thus, multiple lines of evidence indicate that dedicated research to sex, gender, and women’s health is needed to advance health.

Across a variety of diseases, there are sex differences in their underlying biological mechanisms, manifestation, symptomatology, and treatment (11). A lifespan approach is needed as neurodevelopmental disorders, such as autism, are more prevalent in males, and mental health disorders that typically emerge during adolescence and adulthood, such as anxiety and depression, are more prevalent in females (12). In addition, female-specific experiences such as pregnancy, and menopause can impact health outcomes and disease risk (11). For example, hormonal contraceptive use, particularly during adolescence (13) and the postpartum, is associated with increased risk for depression or anxiety disorders (14), suggesting examination of female-specific variables is needed to elucidate mechanisms and new therapeutics for depression. Furthermore, gestational disorders are associated with increased risk for cardiovascular diseases and vascular dementia later in life (15,16), and certain menopause characteristics are linked to an increased risk for CVD and dementia (17,18). These findings suggest that we need to understand why and how these female-specific variables contribute to differences in susceptibility, manifestation and treatment options to reduce burden of disease.

In 2010, the Canadian Institute of Health Research (CIHR) first implemented mandatory fields to indicate whether a project studied and analyzed sex and/or gender as part of their funding application process. In 2019, CIHR introduced grant scoring for sex- and gender-based analysis (SGBA). Analyzing funding trends over time provides insight into what research is prioritized and whether SGBA frameworks introduced by major funding bodies are effective for promoting change. Despite CIHR mandates, under 10% of funded grant abstracts mentioned female, sex, gender, or 2S/LGBTQI health in 2019 and there was a less than 3% increase over the 12 years (2009-2020) examined across all categories (5). Aligned with these findings, a survey of the biomedical literature suggests few publications (5%) are examining sex or gender as a factor in their analyses, despite a greater percentage (68%) of publications reporting on the use of males and females or men and women (19–21).

Examining funding trends over time provides key insight into what research is prioritized and funded. In a previous study, we identified that only 6.6% of CIHR funding identified a female-specific question in their public abstract (5), and under 2% identified examining sex, gender, sexual orientation or gender identity in their abstract. In this study, we wanted to determine if there was a change across time from 2020 (the latest time period previously examined in Stranges et al., 2023) to 2023 in the populations being studied using the funded grant abstracts. Finally, we also analyzed the specific topics of funded female-specific grant abstracts from over 15 years from 2009 to 2023. We were particularly interested in whether funded female-specific CIHR grant abstracts focused on the 11 areas of global disease burden and/or death that disproportionately affect females, and if this has changed over time. We expected that there would be an increase in mention of sex or gender across time since the implementation of SGBA mandates but little change in grants focused on women’s health. We also expected a narrow focus on women’s health topics that would be funded across time.

## Methods

We reviewed publically available abstracts of Project Grants funded by the CIHR in 2021, 2022, and 2023 using the CIHR Funding <u>Decisions Database</u> (22). Project grants are similar to the R01 mechanism at NIH and are open competitions that fund investigator-driven health research projects (https://cihr-irsc.gc.ca/e/49051.html). Prior to 2016, Project Grants were referred to as Operating Grants. All CIHR Operating and Project grant abstracts from 2009-2023 that were awarded during open competitions to Canadian Institutions, regardless of subject population (i.e. human, rodent, cell lines, etc.) were included in our analysis (n=10,629). SGBA was introduced by CIHR in 2010 as a mandatory box for all grant applications and added as a scorable factor for grants in 2019. Thus, this 15 year time span includes grants prior to the introduction of SGBA as well as 4 years post implementation of SGBA as a scorable factor, but 14 years after SGBA was introduced as a mandatory field. This allows us to examine changes in trends after the SGBA mandate on funding topics. We only included funded project or operating grants and not bridge grants (which are awarded for one year only). Grant abstracts in both English and French were examined. French abstracts were translated using the Google translate tool, and then coded.

Funded grant abstracts were coded as “mentioning” using populations that were related to sex (any abstract that would mention sex and/or *males* and *females*, intersex, etc.), gender (any abstract that would mention gender, and/or both *men* and *women*, transgender, non-binary, gender non-conforming, etc.), female-specific (any abstract that would mention only females or *women*), and/or 2S/LGBTQI (any abstract that mentioned members of the 2S/LGBTQI□population specifically or differences in outcomes for 2S/LGBTQI□individuals). This included abstracts that explicitly mentioned matching or controlling for sex or gender. To identify relevant grant abstracts, female-related search terms were used. Female-related search terms were consistent with a previous study (5): female, woman, women, uterus, uter, pregnan, breast, ovary, ovarian, ovaries, girl, menopause, postpartum, maternal, placenta, cervi, vagin, FSF, binary, lesbian, gay, bisexual, queer, LGBT, LGBTQ, LGBTQIA, LGBTQIA□+, LGB, transgender, transsex, trans, 2S, two spirit, 2 spirit, indigi (indigiqueer). Coding was completed by LLG, SAB, GLD, TFLS, and two research assistants and grant abstracts that did not clearly fall into one of these categories were discussed. LLG reviewed all coding. Grant abstracts proposing research across more than one category were double coded. For example, if an abstract mentioned studying lesbian women only it was coded as both female-specific, and 2S/LGBTQI.

A liberal approach was adopted, such that any grant abstracts that mentioned males and females, even though analyzing by sex or sex differences were not explicitly stated, were counted as investigating possible sex differences. For example, an abstract that mentioned using both male and female rodents was coded as “sex”.

To determine what specific areas of female-specific health received more research representation and funding, female-specific abstracts were categorized according to their research area. Of particular interest was whether female-specific abstracts focused on the 11 causes of global disease burden and/or death that have disproportionately affected females over the past three decades (7,8), and whether this focus has shifted over time. Thus, all identified female-specific grants from 2009-2023 (n=681) were coded as to whether they focused on CVDs, cancers, respiratory diseases, diabetes, musculoskeletal disorders, depressive disorders, headache disorders, anxiety, Alzheimer’s disease and other dementias, HIV/AIDS, and low back pain.

### Statistical analyses

To determine whether totals had changed from the last published paper (2020) to 2023, the percentage of funded grants belonging to a given category, as a function of the total number of awarded grants (*n* = 768 from 2023; *n* = 699 from 2020), was calculated. The percentage of funding dollars, as a function of the total funding dollars allocated to the funded grants ($651,948,708 from 2023; $527,933,841 from 2020), was analyzed. All statistical analyses were conducted in RStudio version 4.3.1. Z-tests for proportions using the prop.test() function from the stats package were used to evaluate if there was significant change from 2020 to 2023 in the percentage of grant abstracts that mentioned studying sex, gender, 2S/LGBTQI, were female specific, or did not mention sex/gender as well as the percentage of funding dollars allocated to these same categories. To correct for multiple tests, the p.adjust() function from the stats package was used to apply the false discovery rate (FDR) correction. Cohen’s *h* was calculated as a measure of effect size for each z-test.

To assess what population was examined in grants that mentioned 2S/LGBTQI, a Fisher’s exact test was conducted to compare the proportion of 2S/LGBTQI grants devoted to sexually-/gender-diverse (e.g. gay, queer, bisexual, trans) men and women (23) using the fisher.test() function from the stats package.

Separate analyses were conducted on the female-specific grants from 2009-2023 (*n* = 681). Simple linear regressions were run using the lm() function from the stats package to investigate if there were differences in the percentage of female-specific grants dedicated to the eleven causes of global disease burden that disproportionately affect females, and if there were differences in the percentage of funding dollars dedicated to these causes of global disease burden. Post hoc pairwise comparisons were conducted using the emmeans() function from the emmeans package with Tukey’s adjustment for multiple comparisons, as we were interested in all comparisons. The eff_size() function from the effectsize package was used to obtain Cohen’s *d* for each comparison. Separate linear regressions were used to investigate if the percentage of female-specific grant abstracts mentioning each cause of global disease burden, and the percentage of funding dollars dedicated to each cause of global disease burden, changed over time. To correct for multiple tests, the p.adjust() function from the stats package was used to apply the FDR correction. Statistical significance was set to *p* < 0.05 for all tests. Partial eta squared was calculated as a measure of effect size for each linear model.

## Results

In total, there were 775 project grants that received CIHR funding in 2023. Seven bridge grants were removed, leaving a total of 768 unique project grants that received a total of $651,948,708 in research funding. In 2023, the average project grant received $848,892 in research funding.

### The percentage of grant abstracts mentioning sex or gender doubled in 2023 from 2020 at 5.5% and 1.7% respectively, whereas grant abstracts mentioning 2S/LGBTQI quadrupled during that same time period to 1.8% of the total grant abstracts funded

Compared to 2020, significant improvements were noted in nearly each category of our analysis, with the exception of female health. The percentage of funded grant abstracts that mentioned sex significantly increased, nearly doubling from 2.86% (20 out of 699) in 2020 to 5.47% (42 out of 768) in 2023 (*z* = 2.48, *p* = 0.013, 95% CI: [0.58%, 4.64%]). Similarly, the percentage of grant abstracts that mentioned gender significantly increased, nearly tripling from 0.57% (4 out of 699) in 2020 to 1.69% (13 out of 768) in 2023 (*z* = 2.00, *p* = 0.045, 95% CI: [0.05%, 2.19%]). Moreover, the percentage of grant abstracts that included 2S/LGBTQI significantly increased, quadrupling from 0.43% (3 out of 699) in 2020 to 1.82% (14 out of 768) in 2023 (*z* = 2.49, *p* = 0.013, 95% CI: [0.33%, 2.46%]). Consistent with this, the percentage of grant abstracts that did not mention sex/gender of the populations to be studied significantly decreased from 90.41% (632 out of 699) in 2020 to 85.42% (656 out of 768) in 2023 (*z* = −2.92, *p* = 0.003, 95% CI: [−8.31%, −1.68%]) (Figure 1A).

**Figure 1.**
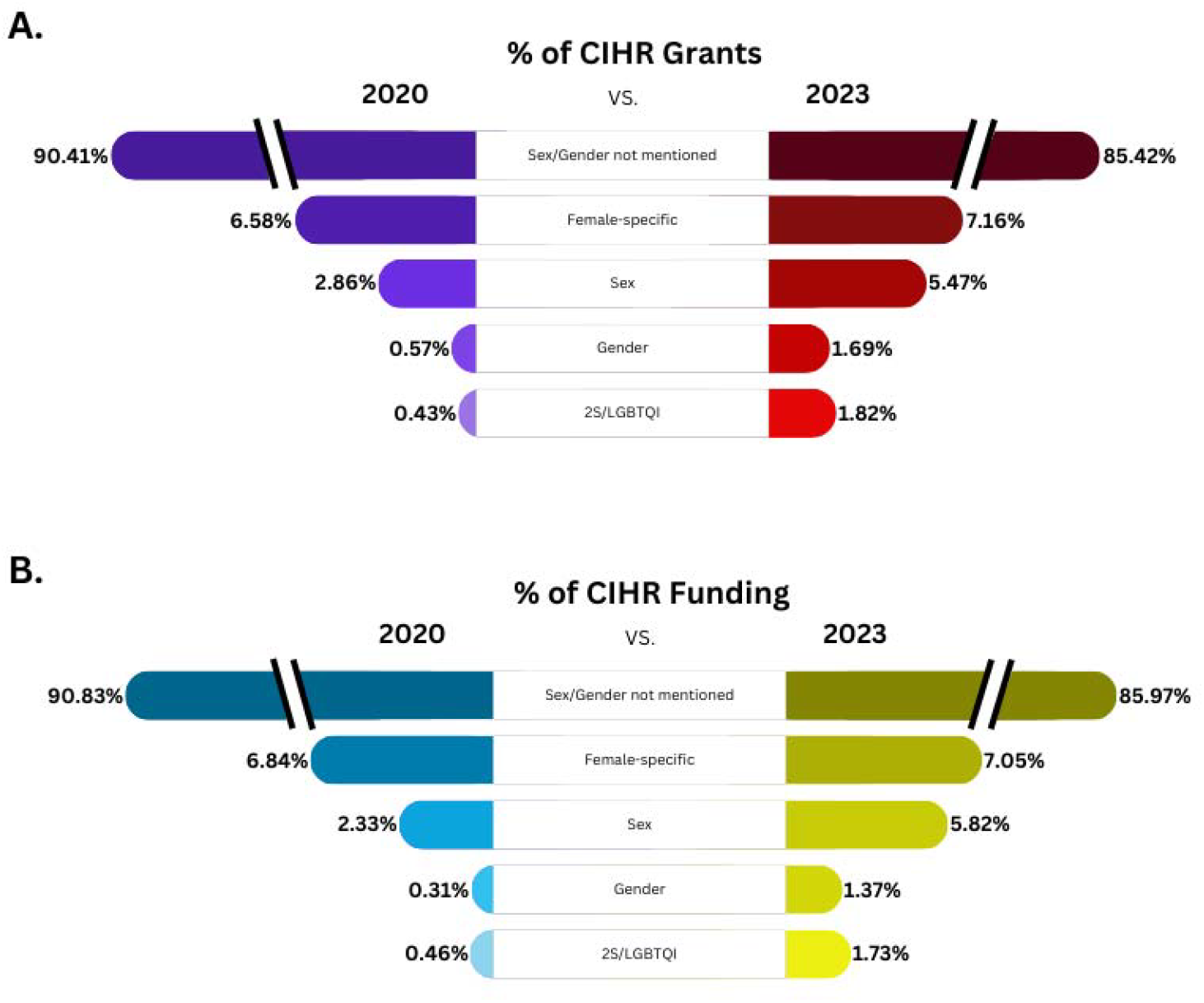
An infographic depicting the change in percentage of grants and funding between 2020 and 2023 for awarded Canadian institutes of Health Research (CIHR) grants for the different categories. The change in percentage (%) of grants (A) and funding amount (B) in the years 2020 and 2023 that did not mention of sex and gender in their grant abstracts or mentioned female-specific health, female-specific health not including cancer based grants, sex, gender, or 2S/LGBTQI health.

### The percentage of grant abstracts mentioning female-specific populations did not significantly change from 2020 to 2023 at 7% of all grant funding

Unlike the increases seen in grant abstracts mentioning sex, gender, or 2S/LGBTQI populations, the percentage of grant abstracts that were female-specific did not significantly increase from 2020 to 2023 (*z* = 0.44, *p* = 0.66, 95% CI: [−2.01%, 3.17%]). Female-specific grant abstracts accounted for 7.16% (55 out of 768) of funded research project grants in 2023, which was a 0.58% increase from 6.58% (46 out of 699) of funded research project grants in 2020.

### Funding totals for grant abstracts mentioning sex or gender doubled in 2023 from 2020, whereas grant abstracts mentioning 2S/LGBTQI quadrupled during that same time period

Increases in funding from 2020 to 2023 amounts largely paralleled findings of representation. The percentage of funding allocated to grants that mentioned sex of the population significantly increased, more than doubling from 2.33% ($12,299,505 out of $527,933,841) in 2020 to 5.82% ($37,935,788 out of $651,948,708) in 2023 (3.49% increase; *z* = 2951.56, *p* < 0.001, 95% CI: [3.47%, 3.49%]). The percentage of funding allocated to grants that mentioned gender significantly increased, quadrupling from 0.31% ($1,625,625 out of $527,933,841) in 2020 to 1.37% ($8,934,047 out of $651,948,708) in 2023 (1.06% increase; *z* = 1926.76, *p* < 0.001, 95% CI: [1.061%, 1.063%]). The percentage of funding allocated to grants that included 2S/LGBTQI populations significantly increased, nearly quadrupling from 0.46% ($2,444,176 out of $527,933,841) in 2020 to 1.73%($11,289,232 out of $651,948,708) in 2023 (1.27% increase; *z* = 2020.18, *p* < 0.001, 95% CI: [1.267%, 1.270%]). Consistent with this, the amount of funding allocated to grant abstracts that did not mention sex/gender significantly decreased from 90.83% ($479,499,892 out of $527,933,841) in 2020 to 85.97% ($560,499,430 out of $651,948,708) in 2023 (4.8% decrease; *z* = −2563.96, *p* < 0.001, 95% CI: [−4.86%, −4.85%]) (Figure 1B).

Although small in magnitude, the percentage of funding allocated to grants that were female-specific significantly increased from 6.84%($36,122,968 out of $527,933,841) in 2020 to 7.05% ($45,965,807 out of $651,948,708) in 2023 (0.21% increase; *z* = 139.76, *p* < 0.001, 95% CI: [0.205%, 0.211%]). When female-specific grant abstracts investigating cancer were removed, the percentage of funding allocated to grants that were female-specific significantly increased from 4.00%($21,096,068 out of $527,933,841) in 2020 to 4.88%($31,817,127 out of $651,948,708) in 2023 (0.88% increase; *z* = 729.79, *p* < 0.001, 95% CI: [0.882%, 0.887%]).

### Twice as many grants that studied S2/LGBTQI persons were focused on male-specific research questions compared to female-specific

Only 1.82% of funded project grants (14 of 768) focused on 2S/LGBTQI persons. Of these, 28.57% focused on sexually/gender-diverse (e.g. gay, queer, bisexual, trans) men and 14.29% focused on sexually/gender-diverse (e.g. lesbian, queer, bisexual, trans) women. Although 2S/LGBTQI research focusing on sexually/gender-diverse men was double that of sexually-/gender-diverse women, the proportion difference was not statistically significant (*OR* = 2.33, *p*-value = 0.65, 95% CI: [0.27 to 30.87]).

### Longitudinal analysis of female-specific grant abstracts

We next examined all the female-specific grant abstracts over the 15 years analyzed (2009-2023) to determine what the research focussed on. There were a total of 681 project grants abstracts that were female-specific out of all 10,629 project grant abstracts (6.49%). These female-specific grant abstracts received 6.40% of total CIHR funding awarded to project/operating grants over the 15 years ($442,179,242 out of $6,910,710,863 in total CIHR funding). We next examined what percentage of female-specific grants mentioned investigating one of the top 11 global burdens of disease and/or death identified for women: CVDs, cancer, diabetes, respiratory diseases, depressive disorders, headache disorders, anxiety, musculoskeletal disorders, Alzheimer’s disease and other dementias, HIV/AIDS, and low back pain (7,8).

### Over 15 years of CIHR funded project abstracts, 4% were dedicated to female-specific project grant abstracts that mentioned the top global burdens of disease and/or death for women

Out of all 681 female-specific grant abstracts spanning 15 years, 482 (70.78%) were dedicated to one of the top 11 global burdens of disease and/or death, which accounted for 4.53% of overall funded grants from CIHR (482 out of 10,629; Figure 2A).

**Figure 2.**
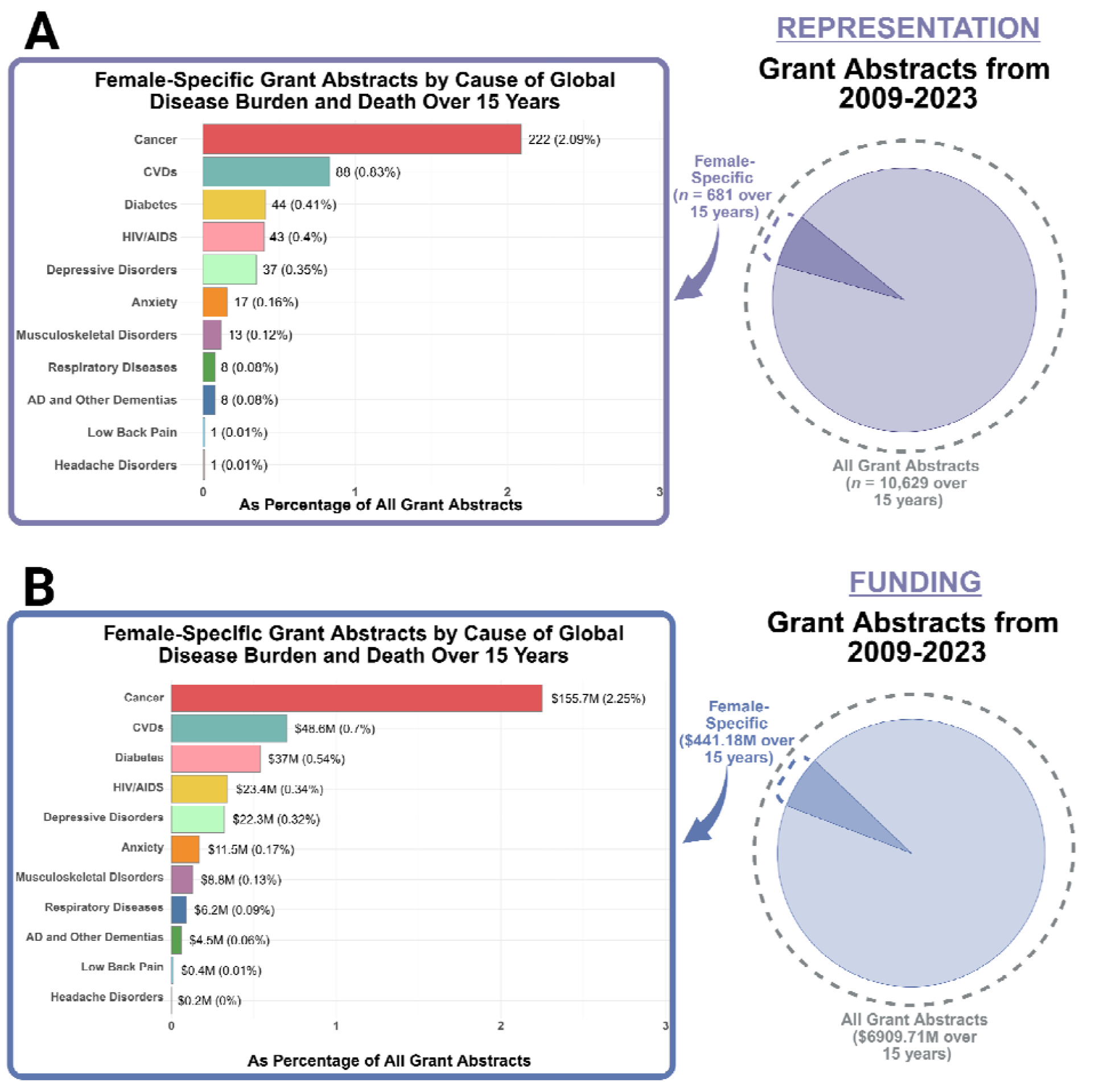
Breakdown of the 681 female-specific grant abstracts from 2009-2023 according to the 11 causes of global disease burden and/or death that disproportionately affect females by A) research representation, and B) funding. Breakdown is provided as a proportion of all grant abstracts. Funding amounts are provided in millions (M). Created in RStudio and https://BioRender.com.

Cancer was the leading category that made up 32.60 % of female-specific grant abstracts (222 of 681) or 2.09% (222 of 10,629) of all funded CIHR grant abstracts over the 15 years.

However, aside from cancer, the 10 other areas of global disease burden received low research representation. Collectively, female-specific grants that mentioned depressive disorders, headache disorders, anxiety, musculoskeletal disorders, Alzheimer’s disease/dementias, HIV/AIDS, CVDs, respiratory diseases, diabetes and low back pain, accounted for 38.18% of female-specific grant abstracts (260 of 681; Figure 2A). In other words, these 10 areas of global disease burden *combined* accounted for 2.45% of the total funded CIHR grant abstracts over the 15 years analyzed (260 of 10,629; Figure 2A) and received approximately the same amount of research representation as cancer.

A linear regression showed that there were significant differences in the percentage of female-specific grant abstracts by cause of global disease burden (*F*(10, 154) =, *p* < 0.001, η^*2*^ = 0.84, 95% CI: [0.80, 1.00]). Post hoc comparisons revealed that the overall percentage of female-specific grant abstracts mentioning cancer was significantly greater than all other causes of global disease burden (for all contrasts with cancer: *t*(154) = 20.14 to 33.01, *p* < 0.0001, Cohen’s *d* = 4.78 to 7.84, see Supplementary Table 1 for all pairwise comparisons). Those mentioning CVDs were also significantly greater than all other causes of global disease burden except cancer (Supplementary Table 1). Additionally, the overall percentage of grant abstracts mentioning low back pain and headache disorders was significantly less than HIV/AIDS, depressive disorders, CVDs, and diabetes (Supplementary Table 1). The overall percentage of grant abstracts mentioning AD and other dementias was significantly less than HIV/AIDS (Supplementary Table 1).

Interestingly, the majority of female-specific grant abstracts that mentioned depressive disorders were dedicated to depressive disorders in pregnancy and postpartum (67.57%; 25 of 37); this is 12.5 times the percentage of grant abstracted dedicated to depressive disorders in perimenopause and menopause (5.41%; 2 of 37). None of the 681 funded female-specific grant abstracts mentioned premenstrual dysphoric disorder (PMDD, 0%) or mentioned studying the relationship between the menstrual cycle and depressive disorders. Similarly, the majority of female-specific grant abstracts that mentioned CVDs were dedicated to CVDs in pregnancy and postpartum (65.91%; 58 of 88); this is nearly 10 times the percentage of grant abstracts dedicated to CVDs in perimenopause and menopause (6.82%; 6 of 88).

Out of the 222 grant female-specific abstracts that mentioned any type of cancer, 23 mentioned more than one type of cancer (17 mentioned two types, and 5 mentioned 3 types, and 1 mentioned 4 types). The overwhelming majority of female-specific abstracts that focused on cancer covered breast (51.19%%; 129 of 252) and gynecologic (43.25%; 109 of 252) cancers (i.e., cervical, endometrial, ovarian, fallopian tube, uterine, or vulvar). The remaining 5.56% covered cancer in general, colorectal, thyroid, throat, skin, liver, oral, ocular, and pancreatic cancers (14 of 252), however, all of these were in combination with breast or gynecologic cancer. In other words, all female-specific grants spanning the 15 years investigating cancer included breast and/or gynecologic cancers as part of their research focus.

### Cancer received a significantly greater percentage of funding than any of the other 10 causes of global disease burden that disproportionately affect females. CVDs and HIV/AIDs also received a significantly greater percentage of funding than most other causes of global disease burden

A linear regression showed that there were significant differences in the percentage of female-specific funding awarded to grant abstracts by cause of global disease burden (*F*(10, 154) = 78.98, *p* < 0.0001, η^*2*^ = 0.84, 95% CI: [0.80, 1.00]; Figure 3B). Post hoc Tukey-adjusted comparisons revealed that the overall percentage of female-specific funding awarded to grant abstracts mentioning cancer was significantly greater than those mentioning the 10 other causes of global disease burden (for all contrasts with cancer: *t*(154) = 24.47 to 35.31, *p* < 0.0001, Cohen’s *d* = 5.56 to 8.03, see Supplementary Table 2 for all pairwise comparisons). The percentage of female-specific funding awarded to grant abstracts mentioning CVDs was significantly greater than all other causes of global disease burden except HIV/AIDS and cancer (Supplementary Table 2). Additionally, female-specific funding awarded to grant abstracts mentioning HIV/AIDS was significantly greater than all other causes of global disease burden except cancer, CVDs, depressive disorders, and diabetes (Supplementary Table 2).

**Figure 3.**
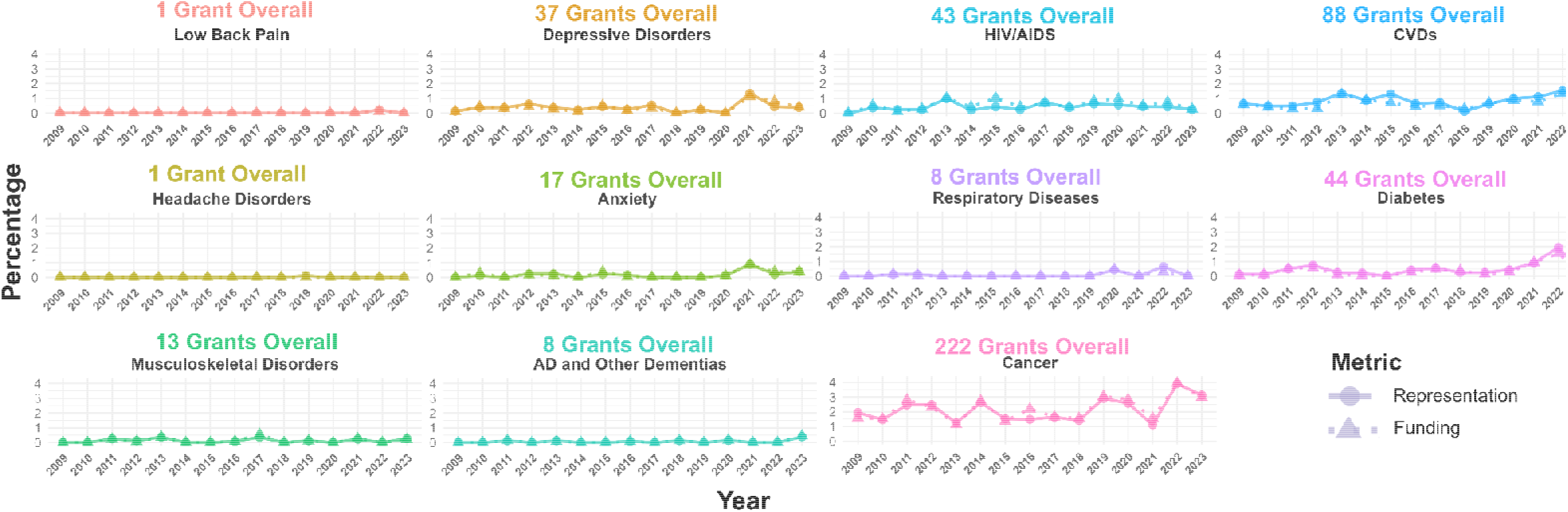
Breakdown of the 681 female-specific grant abstracts over time from 2009-2023 according to the 11 causes of global disease burden and death that disproportionately affect females. Breakdown is provided as a proportion of all funded grant abstracts. Research representation is represented by circles and research funding is represented by triangles. The amount of research representation and funding did not significantly change over time for any of the causes. AD=Alzheimer’s disease, HIV=human immunodeficiency virus, AIDS= Acquired immunodeficiency syndrome, CVDs=cardiovascular diseases. Created in RStudio and https://BioRender.com

### Causes of global disease burden that disproportionately affect females did not show significant changes in representation or funding percentages over time

Linear regressions revealed that the percentage of female-specific grant abstracts mentioning CVDs, anxiety, depressive disorders, musculoskeletal disorders, Alzheimer’s disease and other dementias, HIV/AIDS, headache disorders, low back pain, respiratory diseases, diabetes, or cancer did not show significant changes over time, and the percentage of funding dedicated to these causes of global disease burden also did not significantly change over time (Figure 3; see Supplementary Table 3 for statistics, adjusted *p* ≥ 0.28 for all models).

Lastly we found the percentage of the Canadian women with the top 11 global burden and/or death from disease. Using this we calculated two metrics to show possible disparities between the percentage of funded grant abstracts and the prevalence of these same top 11 global burdens of disease and/or death among Canadian women. We estimated how much more research funding each condition would receive if the funding was proportional to prevalence of that condition in the Canadian female population for comparison purposes. These values are shown in Table 1. Except for funding for HIV/AIDS, the funding is many fold lower than expected based on the prevalence. For example, cancer received only 5% of the expected funding which is 19.5 times lower than expected, and CVD received only 10% of the expected funding which is 10 times lower than expected. The lowest expected funding is based on low back pain and headaches relative to prevalence or both were thousand fold lower than what would be expected, while dementias and respiratory diseases were hundreds times lower than what would be expected based on prevalence.

**Table 1.**
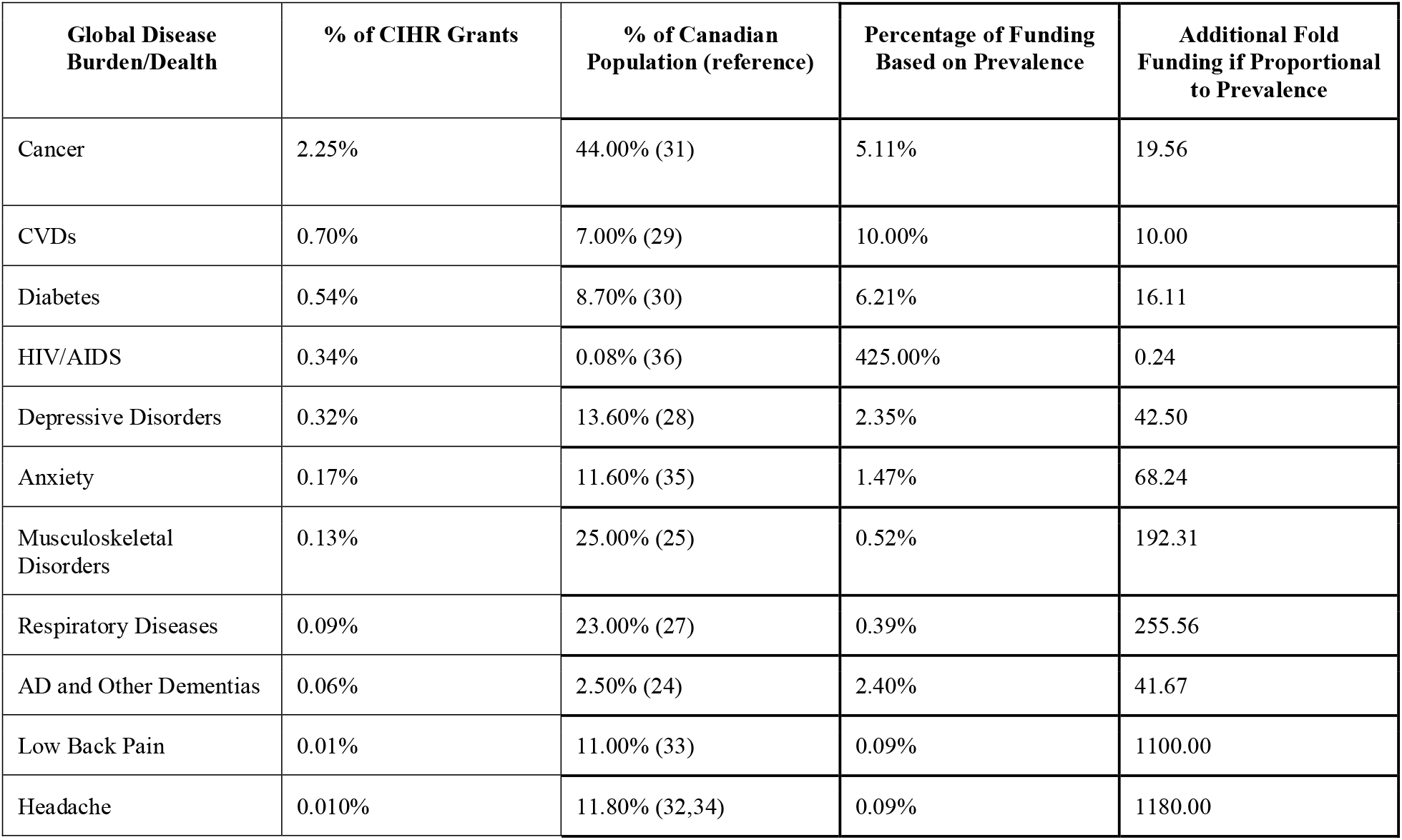
Expected percentage of funding based on Canadian population prevalence for top 11 global burdens of disease and/or death examined. “Percentage of funding based on prevalence” was calculated by dividing the “% of CIHR grants” dedicated to that condition over 15 years by the “% of the Canadian population affected by the condition”. Prevalence data was estimated based on available data, noting that these rates can change across years (e.g. mental health disorders are increasing in Canada). Prevalence was defined usually by lifetime prevalence. CVDs=cardiovascular diseases, HIV=human immunodeficiency virus, AIDS= Acquired immunodeficiency syndrome, AD=Alzheimer’s disease (24–35)

## Discussion

Overall, our analysis of funded CIHR project grant abstracts revealed that the percentage of abstracts mentioning sex or gender doubled and those mentioning 2S/LGBTQI quadrupled from 2020 to 2023. However, despite these gains, the percentages of abstracts mentioning sex, gender or 2S/LGBTQI remained at under 10% of overall funded abstracts. In contrast, progress for female-specific research representation remained unchanged from 2020 to 2023 at ~7%.

Furthermore, the majority of female-specific project grants funded across 15 years of funding from CIHR were focused on breast and gynecological cancers. Cancer research that focused only on females, represented ~2% of total grant abstracts, received significantly more representation and funding than all other causes of global disease burden and/or death that disproportionately affect females. In fact, grant abstracts mentioning the other top 10 causes of global disease burden and/or death *combined* had a similar amount of research representation and funding as cancer research. Indeed, when cancer was excluded, female-specific project grant abstracts mentioning the top 10 global burdens of disease and/or death for women collectively accounted for only 2.45% of all grant abstracts. However, in the Canadian population, cancer is the number 1 cause of death for women, but received 19.5 times lower funding than what would be expected based on prevalence. All other top global burdens and/or death from disease, except for HIV/AIDS, were funded lower than expected which ranged from 10-1800 times lower than expected based on prevalence.

Females are 50.7% of the population in Canada (37), and spend more time in chronic illness than males (3), which underscores the urgent need for increased funding for women’s health research across broad areas of research. Furthermore, no significant increases in funding or research representation dedicated to any of these top causes of disease or death for women were observed over the 15 years. Collectively, our findings suggest that more efforts beyond SGBA+ mandates are needed to increase female-specific research and funding.

### Funded Grant abstracts mentioning studying sex, gender doubled and for 2S/LGBTQI populations quadrupled from 2020 to 2023, but remain under 10% of grants funded

Progress was seen from 2020 to 2023 in grant abstracts mentioning studying sex, gender, or 2S/LGBTQI, but overall increases were small in magnitude. SGBA has been a mandatory inclusion in CIHR grant applications since 2010, it still remains underdisclosed in the public abstracts. This matches surveys of published literature which demonstrates that ~5%-20% of findings, depending on the discipline, analyze with sex/gender as a meaningful variable, rather than using it as a confounder or covariate in the analyses (19,21,38). Reviewers and funding agencies must recognize that there are several types of sex differences beyond more obvious differences in prevalence and risk as diseases can manifest differently between males and females (39). They can be *mechanistic* in nature, such that the cellular machinery underlying a given trait can be different between sexes, or *latent*, such that a sex difference can emerge with a certain condition, stressor, or environment (e.g., age, genotype, disease). Improved education of these types of “hidden” sex differences is important to dispel the myth that female health should solely focus on female reproductive organs.

Relatedly, SGBA requires a more thorough investigation *within* a sex/gender group-especially historically underrepresented groups. Sexual and gender minority groups are more likely to receive delayed breast cancer diagnoses compared to heterosexual women and are 3 times as likely to experience breast cancer recurrence (40,41). These delays in diagnosis and increased rates of recurrence partly stem from a lack of research specific to these groups. Even within 2S/LGBTQI grant abstracts, we found that double the number of abstracts focused on sexually-/gender-diverse men compared to sexually-/gender-diverse women, suggesting that even within gender diverse populations and with sexual orientation research, more research is focused on males. Thus, health knowledge from male only research cannot be generalized to other groups, and inadequate research representation drives gaps in health knowledge and disparities in health outcomes.

### No significant increases in the percentage or amount of funded Women’s Health Research were observed across years. Funding within Women’s Health was narrow and focussed on breast and gynecological cancers

Unlike the increases seen with sex, gender, or 2S/LGBTQI abstracts, there was no significant increase in funding for female-specific populations, which remained at 7%. This suggests that the SGBA mandate is not moving the dial on women’s research. In this study, we also examined what female-specific research was funded across 681grant abstracts across 15 years using the top 11 global burden of disease and death for women as a guide. Our analyses revealed that cancer received significantly more research representation and funding, approximately the same amount of research representation as the 10 other global burdens of disease and death *combined*. Yet still, the amount of funding dedicated to cancer in females was 19 fold less than what might be expected based on the population of Canadian women affected.

Moreover, even within the field of cancer, all funded cancer-related female-specific project grants included breast and gynecologic cancers. In fact, only 5.56% of cancer-related female-specific project grants additionally included another type of cancer (e.g., colorectal, thyroid, pancreatic). It is imperative to research breast and gynecological cancers, which account for approximately 36.4% of cancers in women in Canada (26). However, lung cancer is the second most common cancer in Canadian women, and is more frequently diagnosed in women than men, but it was not represented in funded grants from 2009-2023, suggesting other cancer types are limited from research (26). Thus, our analysis revealed that the breadth of female-specific research is narrow.

### Of the remaining top 10 global burden and/or death of disease for women besides cancer, CIHR funded research accounted for less than 3%

Of the other top 10 global burden and/or death from disease in women beyond cancer, funding representation was very low at 2.45%. As seen in Table 1, there was a disproportionate representation of funding for the top global burden and/or death compared to the Canadian population except for HIV/AIDS which was disproportionately higher than the population affected. Although most of the top 11 global burden and/or death of diseases were disproportionately under-represented in CIHR funding compared to the expected Canadian prevalence, the areas that were over 30x less likely to be funded were headache disorders, low back pain, respiratory diseases, dementia, anxiety and depressive disorders. Female-specific factors such menstrual cycles, pregnancy, and menopause contribute to the greater risk for dementia, migraines and depression/anxiety among females (17,42,43). More studies are needed to uncover why these factors confer a greater risk to develop personalized treatments.Headache disorders were the lowest proportionately funded compared to the affected Canadian population (32,44), one type of headache disorder, affect approximately 11-20% of Canadian women (32,45) *one* female-specific funded grant abstract was dedicated to headache disorders in women over 15 years. Low back pain, which disproportionately affects women and approximately 619 million people globally and 11% of Canadian women (33,46) was also only mentioned once in any female-specific funded grant abstract over 15 years. Respiratory diseases, such asthma, were disproportionately represented compared to the Canadian population at > 200x. Approximately 6% of Canadian women are diagnosed with dementia yet CIHR funding represented only 1% of the affected female population, representing a 1/100th of the expected funding based on prevalence. In Canada, 13% of the female population will experience depression (28) and 11% will experience anxiety disorders (35), however, only 0.37% of funded grants were for female-specific studies on anxiety and depression over 15 years, indicating a disproportionate proportion of funding. Furthermore, many of these conditions were largely studied in the context of pregnancy. None of the 681 funded female-specific grant abstracts over 15 years mentioned premenstrual dysphoric disorder, or the relationship between the menstrual cycle and depressive disorders. Given the potential link of mental health conditions, including anxiety and depression, to ovarian hormones, there is a need to investigate the influence of various hormonal transitions across the lifespan on the brain.

### Limitations

One limitation is that we were restricted to publicly available data, which is currently confined to the public abstract, which has word constraints. Thus, it is possible that research integrating SGBA, specific populations of interest, or diseases/conditions of interest were missed due to the word constraints. However, our bar for categorization was low, such that if an abstract simply mentioned including the population/disease/condition of interest, it was sufficient to be categorized as such. Relatedly, we were unable to assess if optimal statistical analyses were appropriately selected to meaningfully uncover sex or gender differences, e.g., avoiding sex or gender as covariates, and avoiding using matched samples. In an in depth analysis of published psychiatry and neuroscience research articles, optimal statistical analyses are often not selected (21), which is similar to findings across multiple disciplines (19,20).

Although not the focus of our study, it is also possible that males are ignored in research for diseases and conditions that affect all sexes, but are more prevalent in females. We observed that out of the 26 project grants from 2023 that focused on breast cancer, none focused exclusively on breast cancer in males. Autoimmune conditions, osteoporosis, anxiety and depression disproportionately affect females, but still influence males and intersex individuals. We recognize that research on these diseases and conditions in non-female populations is also important, and is an area that should be explored in future research. However, recognizing that female-specific research continues to be underrepresented - especially in the top causes of global disease burden and death - suggests that efforts beyond SGBA mandates are needed to increase female-specific research funding.

### A path forward: Recommendations beyond mandates

Our analysis suggests that more SGBA efforts, beyond mandates, are necessary to increase research on sex, gender, 2S/LGBTQI, and most notably female-specific research. Female-specific research was the only area in our analysis that did not show an increase in research representation across the 15 years. Females make up approximately 50% of the population, yet research pertaining to females only received only 7% of all CIHR project grant research representation and funding.

Dedicated research funding is essential for advancing women’s health and furthering knowledge generation. Examples of dedicated research funding for women’s health research come from the Gates Foundation (47), the Canadian Heart and Stroke Foundation Women’s Research Network (48) and Wellcome Leap’s Cutting Alzheimer’s Risk through Endocrinology (CARE) grant (49). These funding announcements are steps in the right direction for closing the women’s health gap. However, protected funding is needed, especially for the 11 causes of disease burden and/or death that disproportionately affect females.

It is not clear why SGBA has not led to an increase in women’s health research funding but it may be related to a lack of awareness and opportunities for women’s research topics.

Having more women’s health special issues in renowned health journals is critical to closing the women’s health gap. These special issues reinforce the value of women’s health research and provide an opportunity to further women’s health knowledge in areas beyond reproductive health (i.e. health related to reproductive organs (breast, ovaries), fertility, sexual health, and/or pregnancy). The Journal of the American Medical Association (JAMA) International Medicine series “Improving Women’s Health Across the Life Span” was introduced in 2024 to increase published articles relating to women’s health (50). Moreover, closing the women’s health gap would have significant global economic benefits, boosting the global economy by at least $1 trillion annually by 2040 (3).

## Conclusions

A major strength of our study is the longitudinal analysis, which was conducted using data from 15 years of CIHR abstracts. This longitudinal approach provided valuable insights into trends in research representation and funding over time. Moreover, a detailed thematic analysis of the female-specific grant abstracts provided further insight into their content, and revealed gaps in the focus of the grant abstracts relative to the prevalence of diseases in the Canadian population; it revealed a significant emphasis on pregnancy research compared to any other time point in a female’s lifespan.

Female health research did not increase over time and continues to be significantly lacking in areas outside of reproductive health as well as toward the top female causes of global disease burden and death. These gaps are even greater for marginalized groups, including women of colour and women with disabilities (51,52). Dedicated, protected research funding for women’s health and women’s health special issues in top health journals is essential to diversify female-specific research and advance basic science centered on women. Critically, researching women’s health factors will have significant economic benefits and will improve personalized medicine and health outcomes for everyone, not just women.

## Data Availability

All data produced in the present study are available upon reasonable request to the authors

## Declarations

### Ethics Approval and Consent to Participate

Not applicable

## Consent for Publication

Not applicable

## Availability of Data and Materials Availability

All material was drawn from publicly available databases but is available upon reasonable request.

## Authors’ Contributions

L.L.G. and L.A.M.G. designed the study and were responsible for the methodology. L.L.G. was responsible for formal analysis, data visualisation, and wrote the original draft of the manuscript. L.L.G., T.F.L.S., A.M., S.A.B., and G.L.D. contributed to data curation, including data coding.

L.A.M.G. was responsible for funding acquisition and study supervision. All authors reviewed and edited the manuscript.

## Acknowledgements

We are deeply grateful to Tori N. Stranges and Amanda B. Namchuk for their invaluable contributions in coding abstracts from 2009-2020, as well as Evita Jiang and Yagoda Oleksak for their assistance with coding abstracts from 2021 and 2022.

