## Supplement for "Women’s Health Research Funding in Canada across 15 years suggests low funding levels with a narrow focus"

**Supplementary Material**

**Table S1**

| **Contrast** | **Estimate** | ***SE*** | ***df*** | ***t*-ratio** | ***p*-value** | **Cohen’s *d*** |
| --- | --- | --- | --- | --- | --- | --- |
| **(HIV/AIDS) - Low Back Pain** | 6.18 | 1.54 | 154 | 4.02 | 0.00 | 1.47 |
| **(HIV/AIDS) - Musculoskeletal Disorders** | 4.58 | 1.54 | 154 | 2.98 | 0.11 | 1.09 |
| **(HIV/AIDS) - Respiratory Diseases** | 5.26 | 1.54 | 154 | 3.42 | 0.03 | 1.25 |
| **AD and Other Dementias - (HIV/AIDS)** | -5.20 | 1.54 | 154 | -3.38 | 0.04 | -1.23 |
| **AD and Other Dementias - Anxiety** | -1.20 | 1.54 | 154 | -0.78 | 1.00 | -0.28 |
| **AD and Other Dementias - Cancer** | -32.03 | 1.54 | 154 | -20.83 | 0.00 | -7.61 |
| **AD and Other Dementias - CVDs** | -11.89 | 1.54 | 154 | -7.73 | 0.00 | -2.82 |
| **AD and Other Dementias - Depressive Disorders** | -4.25 | 1.54 | 154 | -2.77 | 0.18 | -1.01 |
| **AD and Other Dementias - Diabetes** | -4.89 | 1.54 | 154 | -3.18 | 0.06 | -1.16 |
| **AD and Other Dementias - Headache Disorders** | 0.97 | 1.54 | 154 | 0.63 | 1.00 | 0.23 |
| **AD and Other Dementias - Low Back Pain** | 0.98 | 1.54 | 154 | 0.64 | 1.00 | 0.23 |
| **AD and Other Dementias - Musculoskeletal Disorders** | -0.62 | 1.54 | 154 | -0.40 | 1.00 | -0.15 |
| **AD and Other Dementias - Respiratory Diseases** | 0.06 | 1.54 | 154 | 0.04 | 1.00 | 0.01 |
| **Anxiety - (HIV/AIDS)** | -4.00 | 1.54 | 154 | -2.60 | 0.26 | -0.95 |
| **Anxiety - Cancer** | -30.83 | 1.54 | 154 | -20.05 | 0.00 | -7.32 |
| **Anxiety - CVDs** | -10.69 | 1.54 | 154 | -6.95 | 0.00 | -2.54 |
| **Anxiety - Depressive Disorders** | -3.05 | 1.54 | 154 | -1.99 | 0.66 | -0.73 |
| **Anxiety - Diabetes** | -3.69 | 1.54 | 154 | -2.40 | 0.37 | -0.88 |
| **Anxiety - Headache Disorders** | 2.17 | 1.54 | 154 | 1.41 | 0.94 | 0.52 |
| **Anxiety - Low Back Pain** | 2.18 | 1.54 | 154 | 1.42 | 0.94 | 0.52 |
| **Anxiety - Musculoskeletal Disorders** | 0.58 | 1.54 | 154 | 0.38 | 1.00 | 0.14 |
| **Anxiety - Respiratory Diseases** | 1.26 | 1.54 | 154 | 0.82 | 1.00 | 0.30 |
| **Cancer - (HIV/AIDS)** | 26.83 | 1.54 | 154 | 17.45 | 0.00 | 6.37 |
| **Cancer - CVDs** | 20.14 | 1.54 | 154 | 13.10 | 0.00 | 4.78 |
| **Cancer - Depressive Disorders** | 27.78 | 1.54 | 154 | 18.07 | 0.00 | 6.60 |
| **Cancer - Diabetes** | 27.14 | 1.54 | 154 | 17.65 | 0.00 | 6.45 |
| **Cancer - Headache Disorders** | 33.00 | 1.54 | 154 | 21.46 | 0.00 | 7.84 |
| **Cancer - Low Back Pain** | 33.01 | 1.54 | 154 | 21.47 | 0.00 | 7.84 |
| **Cancer - Musculoskeletal Disorders** | 31.41 | 1.54 | 154 | 20.43 | 0.00 | 7.46 |
| **Cancer - Respiratory Diseases** | 32.09 | 1.54 | 154 | 20.87 | 0.00 | 7.62 |
| **CVDs - (HIV/AIDS)** | 6.69 | 1.54 | 154 | 4.35 | 0.00 | 1.59 |
| **CVDs - Depressive Disorders** | 7.63 | 1.54 | 154 | 4.96 | 0.00 | 1.81 |
| **CVDs - Diabetes** | 7.00 | 1.54 | 154 | 4.55 | 0.00 | 1.66 |
| **CVDs - Headache Disorders** | 12.86 | 1.54 | 154 | 8.36 | 0.00 | 3.05 |
| **CVDs - Low Back Pain** | 12.87 | 1.54 | 154 | 8.37 | 0.00 | 3.06 |
| **CVDs - Musculoskeletal Disorders** | 11.27 | 1.54 | 154 | 7.33 | 0.00 | 2.68 |
| **CVDs - Respiratory Diseases** | 11.95 | 1.54 | 154 | 7.77 | 0.00 | 2.84 |
| **Depressive Disorders - (HIV/AIDS)** | -0.94 | 1.54 | 154 | -0.61 | 1.00 | -0.22 |
| **Depressive Disorders - Diabetes** | -0.64 | 1.54 | 154 | -0.41 | 1.00 | -0.15 |
| **Depressive Disorders - Headache Disorders** | 5.22 | 1.54 | 154 | 3.40 | 0.03 | 1.24 |
| **Depressive Disorders - Low Back Pain** | 5.23 | 1.54 | 154 | 3.40 | 0.03 | 1.24 |
| **Depressive Disorders - Musculoskeletal Disorders** | 3.63 | 1.54 | 154 | 2.36 | 0.40 | 0.86 |
| **Depressive Disorders - Respiratory Diseases** | 4.31 | 1.54 | 154 | 2.81 | 0.17 | 1.02 |
| **Diabetes - (HIV/AIDS)** | -0.31 | 1.54 | 154 | -0.20 | 1.00 | -0.07 |
| **Diabetes - Headache Disorders** | 5.86 | 1.54 | 154 | 3.81 | 0.01 | 1.39 |
| **Diabetes - Low Back Pain** | 5.87 | 1.54 | 154 | 3.82 | 0.01 | 1.39 |
| **Diabetes - Musculoskeletal Disorders** | 4.27 | 1.54 | 154 | 2.78 | 0.18 | 1.01 |
| **Diabetes - Respiratory Diseases** | 4.95 | 1.54 | 154 | 3.22 | 0.06 | 1.18 |
| **Headache Disorders - (HIV/AIDS)** | -6.17 | 1.54 | 154 | -4.01 | 0.00 | -1.46 |
| **Headache Disorders - Low Back Pain** | 0.01 | 1.54 | 154 | 0.01 | 1.00 | 0.00 |
| **Headache Disorders - Musculoskeletal Disorders** | -1.59 | 1.54 | 154 | -1.03 | 0.99 | -0.38 |
| **Headache Disorders - Respiratory Diseases** | -0.91 | 1.54 | 154 | -0.59 | 1.00 | -0.22 |
| **Low Back Pain - Musculoskeletal Disorders** | -1.60 | 1.54 | 154 | -1.04 | 0.99 | -0.38 |
| **Low Back Pain - Respiratory Diseases** | -0.92 | 1.54 | 154 | -0.60 | 1.00 | -0.22 |
| **Musculoskeletal Disorders - Respiratory Diseases** | 0.68 | 1.54 | 154 | 0.44 | 1.00 | 0.16 |

**Table S2**

| **Contrast** | **Estimate** | ***SE*** | ***df*** | ***t*-ratio** | ***p*-value** | **Cohen’s *d*** |
| --- | --- | --- | --- | --- | --- | --- |
| **(HIV/AIDS) - Low Back Pain** | 8.48 | 1.61 | 154 | 5.28 | 0.00 | 1.93 |
| **(HIV/AIDS) - Musculoskeletal Disorders** | 6.63 | 1.61 | 154 | 4.13 | 0.00 | 1.51 |
| **(HIV/AIDS) - Respiratory Diseases** | 7.69 | 1.61 | 154 | 4.79 | 0.00 | 1.75 |
| **AD and Other Dementias - (HIV/AIDS)** | -7.39 | 1.61 | 154 | -4.60 | 0.00 | -1.68 |
| **AD and Other Dementias - Anxiety** | -1.24 | 1.61 | 154 | -0.77 | 1.00 | -0.28 |
| **AD and Other Dementias - Cancer** | -34.18 | 1.61 | 154 | -21.28 | 0.00 | -7.77 |
| **AD and Other Dementias - CVDs** | -9.71 | 1.61 | 154 | -6.05 | 0.00 | -2.21 |
| **AD and Other Dementias - Depressive Disorders** | -3.82 | 1.61 | 154 | -2.38 | 0.39 | -0.87 |
| **AD and Other Dementias - Diabetes** | -3.79 | 1.61 | 154 | -2.36 | 0.40 | -0.86 |
| **AD and Other Dementias - Headache Disorders** | 1.13 | 1.61 | 154 | 0.70 | 1.00 | 0.26 |
| **AD and Other Dementias - Low Back Pain** | 1.09 | 1.61 | 154 | 0.68 | 1.00 | 0.25 |
| **AD and Other Dementias - Musculoskeletal Disorders** | -0.77 | 1.61 | 154 | -0.48 | 1.00 | -0.17 |
| **AD and Other Dementias - Respiratory Diseases** | 0.29 | 1.61 | 154 | 0.18 | 1.00 | 0.07 |
| **Anxiety - (HIV/AIDS)** | -6.16 | 1.61 | 154 | -3.83 | 0.01 | -1.40 |
| **Anxiety - Cancer** | -32.95 | 1.61 | 154 | -20.51 | 0.00 | -7.49 |
| **Anxiety - CVDs** | -8.47 | 1.61 | 154 | -5.27 | 0.00 | -1.93 |
| **Anxiety - Depressive Disorders** | -2.58 | 1.61 | 154 | -1.61 | 0.88 | -0.59 |
| **Anxiety - Diabetes** | -2.55 | 1.61 | 154 | -1.59 | 0.89 | -0.58 |
| **Anxiety - Headache Disorders** | 2.37 | 1.61 | 154 | 1.47 | 0.93 | 0.54 |
| **Anxiety - Low Back Pain** | 2.33 | 1.61 | 154 | 1.45 | 0.93 | 0.53 |
| **Anxiety - Musculoskeletal Disorders** | 0.47 | 1.61 | 154 | 0.29 | 1.00 | 0.11 |
| **Anxiety - Respiratory Diseases** | 1.53 | 1.61 | 154 | 0.95 | 1.00 | 0.35 |
| **Cancer - (HIV/AIDS)** | 26.79 | 1.61 | 154 | 16.68 | 0.00 | 6.09 |
| **Cancer - CVDs** | 24.47 | 1.61 | 154 | 15.24 | 0.00 | 5.56 |
| **Cancer - Depressive Disorders** | 30.36 | 1.61 | 154 | 18.90 | 0.00 | 6.90 |
| **Cancer - Diabetes** | 30.40 | 1.61 | 154 | 18.93 | 0.00 | 6.91 |
| **Cancer - Headache Disorders** | 35.31 | 1.61 | 154 | 21.99 | 0.00 | 8.03 |
| **Cancer - Low Back Pain** | 35.27 | 1.61 | 154 | 21.96 | 0.00 | 8.02 |
| **Cancer - Musculoskeletal Disorders** | 33.42 | 1.61 | 154 | 20.81 | 0.00 | 7.60 |
| **Cancer - Respiratory Diseases** | 34.48 | 1.61 | 154 | 21.47 | 0.00 | 7.84 |
| **CVDs - (HIV/AIDS)** | 2.32 | 1.61 | 154 | 1.44 | 0.94 | 0.53 |
| **CVDs - Depressive Disorders** | 5.89 | 1.61 | 154 | 3.67 | 0.01 | 1.34 |
| **CVDs - Diabetes** | 5.93 | 1.61 | 154 | 3.69 | 0.01 | 1.35 |
| **CVDs - Headache Disorders** | 10.84 | 1.61 | 154 | 6.75 | 0.00 | 2.46 |
| **CVDs - Low Back Pain** | 10.80 | 1.61 | 154 | 6.72 | 0.00 | 2.45 |
| **CVDs - Musculoskeletal Disorders** | 8.94 | 1.61 | 154 | 5.57 | 0.00 | 2.03 |
| **CVDs - Respiratory Diseases** | 10.00 | 1.61 | 154 | 6.23 | 0.00 | 2.27 |
| **Depressive Disorders - (HIV/AIDS)** | -3.57 | 1.61 | 154 | -2.23 | 0.49 | -0.81 |
| **Depressive Disorders - Diabetes** | 0.03 | 1.61 | 154 | 0.02 | 1.00 | 0.01 |
| **Depressive Disorders - Headache Disorders** | 4.95 | 1.61 | 154 | 3.08 | 0.08 | 1.12 |
| **Depressive Disorders - Low Back Pain** | 4.91 | 1.61 | 154 | 3.06 | 0.09 | 1.12 |
| **Depressive Disorders - Musculoskeletal Disorders** | 3.05 | 1.61 | 154 | 1.90 | 0.72 | 0.69 |
| **Depressive Disorders - Respiratory Diseases** | 4.11 | 1.61 | 154 | 2.56 | 0.28 | 0.94 |
| **Diabetes - (HIV/AIDS)** | -3.61 | 1.61 | 154 | -2.25 | 0.48 | -0.82 |
| **Diabetes - Headache Disorders** | 4.91 | 1.61 | 154 | 3.06 | 0.09 | 1.12 |
| **Diabetes - Low Back Pain** | 4.87 | 1.61 | 154 | 3.03 | 0.09 | 1.11 |
| **Diabetes - Musculoskeletal Disorders** | 3.02 | 1.61 | 154 | 1.88 | 0.73 | 0.69 |
| **Diabetes - Respiratory Diseases** | 4.08 | 1.61 | 154 | 2.54 | 0.29 | 0.93 |
| **Headache Disorders - (HIV/AIDS)** | -8.52 | 1.61 | 154 | -5.31 | 0.00 | -1.94 |
| **Headache Disorders - Low Back Pain** | -0.04 | 1.61 | 154 | -0.03 | 1.00 | -0.01 |
| **Headache Disorders - Musculoskeletal Disorders** | -1.89 | 1.61 | 154 | -1.18 | 0.98 | -0.43 |
| **Headache Disorders - Respiratory Diseases** | -0.83 | 1.61 | 154 | -0.52 | 1.00 | -0.19 |
| **Low Back Pain - Musculoskeletal Disorders** | -1.85 | 1.61 | 154 | -1.15 | 0.99 | -0.42 |
| **Low Back Pain - Respiratory Diseases** | -0.79 | 1.61 | 154 | -0.49 | 1.00 | -0.18 |
| **Musculoskeletal Disorders - Respiratory Diseases** | 1.06 | 1.61 | 154 | 0.66 | 1.00 | 0.24 |
| **(HIV/AIDS) - Low Back Pain** | 8.48 | 1.61 | 154 | 5.28 | 0.00 | 1.93 |
| **(HIV/AIDS) - Musculoskeletal Disorders** | 6.63 | 1.61 | 154 | 4.13 | 0.00 | 1.51 |
| **(HIV/AIDS) - Respiratory Diseases** | 7.69 | 1.61 | 154 | 4.79 | 0.00 | 1.75 |

**Table S3**

| **Category** | **Term** | **Estimate** | ***SE*** | ***t-*statistic** | ***p-*value** | **Adjusted *p-*value** | ***η²*** |
| --- | --- | --- | --- | --- | --- | --- | --- |
| HIV/AIDS | as.numeric(year) | 0.01 | 0.22 | 0.05 | 0.96 | 0.99 | 0.00 |
| Anxiety | as.numeric(year) | 0.16 | 0.14 | 1.18 | 0.26 | 0.70 | 0.10 |
| Depression | as.numeric(year) | -0.16 | 0.19 | -0.82 | 0.43 | 0.70 | 0.05 |
| AD and Other Dementias | as.numeric(year) | 0.13 | 0.09 | 1.41 | 0.18 | 0.70 | 0.13 |
| Musculoskeletal Disorders | as.numeric(year) | 0.00 | 0.12 | -0.01 | 0.99 | 0.99 | 0.00 |
| Headache Disorders | as.numeric(year) | 0.02 | 0.03 | 0.68 | 0.51 | 0.70 | 0.03 |
| Respiratory Diseases | as.numeric(year) | 0.13 | 0.12 | 1.09 | 0.30 | 0.70 | 0.08 |
| CVDs | as.numeric(year) | -0.05 | 0.31 | -0.17 | 0.87 | 0.99 | 0.00 |
| Diabetes | as.numeric(year) | 0.17 | 0.24 | 0.69 | 0.50 | 0.70 | 0.04 |
| Cancer | as.numeric(year) | -0.57 | 0.65 | -0.89 | 0.39 | 0.70 | 0.06 |
| Low Back Pain | as.numeric(year) | 0.03 | 0.02 | 1.44 | 0.17 | 0.70 | 0.14 |
| HIV/AIDS Funding | as.numeric(year) | 0.08 | 0.31 | 0.27 | 0.80 | 0.97 | 0.01 |
| Anxiety Funding | as.numeric(year) | 0.15 | 0.16 | 0.92 | 0.37 | 0.80 | 0.06 |
| Depression Funding | as.numeric(year) | -0.08 | 0.18 | -0.44 | 0.67 | 0.92 | 0.01 |
| AD and Other Dementias Funding | as.numeric(year) | 0.16 | 0.10 | 1.54 | 0.15 | 0.80 | 0.15 |
| Musculoskeletal Disorders Funding | as.numeric(year) | 0.00 | 0.16 | 0.02 | 0.98 | 0.98 | 0.00 |
| Headache Disorders Funding | as.numeric(year) | 0.00 | 0.01 | 0.68 | 0.51 | 0.80 | 0.03 |
| Respiratory Diseases Funding | as.numeric(year) | 0.09 | 0.11 | 0.81 | 0.43 | 0.80 | 0.05 |
| CVDs Funding | as.numeric(year) | 0.02 | 0.27 | 0.08 | 0.94 | 0.98 | 0.00 |
| Diabetes Funding | as.numeric(year) | 0.18 | 0.25 | 0.73 | 0.48 | 0.80 | 0.04 |
| Cancer Funding | as.numeric(year) | -0.51 | 0.67 | -0.76 | 0.46 | 0.80 | 0.04 |
| Low Back Pain Funding | as.numeric(year) | 0.02 | 0.01 | 1.44 | 0.17 | 0.80 | 0.14 |
